# Peripheral neural synchrony in post-lingually deafened adult cochlear implant users

**DOI:** 10.1101/2023.07.07.23292369

**Authors:** Shuman He, Jeffrey Skidmore, Ian C. Bruce, Jacob J. Oleson, Yi Yuan

## Abstract

**Objective:** This paper reports a noninvasive method for quantifying neural synchrony in the cochlear nerve (i.e., peripheral neural synchrony) in cochlear implant (CI) users, which allows for evaluating this physiological phenomenon in human CI users for the first time in the literature. In addition, this study assessed how peripheral neural synchrony was correlated with temporal resolution acuity and speech perception outcomes measured in quiet and in noise in post-lingually deafened adult CI users. It tested the hypothesis that peripheral neural synchrony was an important factor for temporal resolution acuity and speech perception outcomes in noise in post-lingually deafened adult CI users.

**Design:** Study participants included 24 post-lingually deafened adult CI users with a Cochlear™ Nucleus® device. Three study participants were implanted bilaterally, and each ear was tested separately. For each of the 27 implanted ears tested in this study, 400 sweeps of the electrically evoked compound action potential (eCAP) were measured at four electrode locations across the electrode array. Peripheral neural synchrony was quantified at each electrode location using the phase locking value (PLV), which is a measure of trial-by-trial phase coherence among eCAP sweeps/trials. Temporal resolution acuity was evaluated by measuring the within-channel gap detection threshold (GDT) using a three-alternative, forced-choice procedure in a subgroup of 20 participants (23 implanted ears). For each ear tested in these participants, GDTs were measured at two electrode locations with a large difference in PLVs. For 26 implanted ears tested in 23 participants, speech perception performance was evaluated using Consonant-Nucleus-Consonant (CNC) word lists presented in quiet and in noise at signal-to-noise ratios (SNRs) of +10 and +5 dB. Linear Mixed effect Models were used to evaluate the effect of electrode location on the PLV and the effect of the PLV on GDT after controlling for the stimulation level effects. Pearson product-moment correlation tests were used to assess the correlations between PLVs, CNC word scores measured in different conditions, and the degree of noise effect on CNC word scores.

**Results:** There was a significant effect of electrode location on the PLV after controlling for the effect of stimulation level. There was a significant effect of the PLV on GDT after controlling for the effects of stimulation level, where higher PLVs (greater synchrony) led to lower GDTs (better temporal resolution acuity). PLVs were not significantly correlated with CNC word scores measured in any listening condition or the effect of competing background noise presented at a SNR of +10 dB on CNC word scores. In contrast, there was a significant negative correlation between the PLV and the degree of noise effect on CNC word scores for a competing background noise presented at a SNR of +5 dB, where higher PLVs (greater synchrony) correlated with smaller noise effects on CNC word scores.

**Conclusions:** This newly developed method can be used to assess peripheral neural synchrony in CI users, a physiological phenomenon that has not been systematically evaluated in electrical hearing. Poorer peripheral neural synchrony leads to lower temporal resolution acuity and is correlated with a larger detrimental effect of competing background noise presented at a SNR of 5 dB on speech perception performance in post-lingually deafened adult CI users.

## INTRODUCTION

While many cochlear implant (CI) users can achieve excellent listening outcomes in quiet, speech recognition in background noise remains a significant challenge (Eisenberg et al., 2016; Torkildsen et al., 2019; Zaltz et al., 2020). The neural mechanisms underlying the observed speech perception deficits in noise in CI users remain unknown. In acoustic hearing, discharge synchronization of cochlear nerve (CN) fibers has been shown to play a critical role in neural representation of speech sounds presented in noise in animal models (e.g., Delgutte & Kiang, 1984; Heeringa & Koppl, 2022; Sachs et al., 1983). Simulation results from computational models demonstrated the importance of synchronized neural firing from CN fibers for robust encoding of consonants in spectro-temporally modulated background noises (Bruce et al., 2013; Viswanathan et al., 2022). These simulation results also showed that poor neural synchrony in the CN (i.e., peripheral neural synchrony) results in smeared neural representation of temporal envelope cues, which leads to deficits in processing these cues (Zeng et al., 2005; Zeng et al., 1999). Aligned with these results from animal models and computational simulations, listeners with poor peripheral neural synchrony (e.g., patients with auditory neuropathy spectrum disorder and elderly listeners) have temporal processing deficits and show excessive difficulties in understanding speech in noise (e.g., Harris et al., 2021; Kraus et al., 2000; Rance, 2005; Zeng et al., 2005). Overall, these results demonstrate the importance of peripheral neural synchrony for temporal processing and speech perception in noise in acoustic hearing.

Deteriorations in anatomical structures of the CN in CI patients have been well established based on the histological results of human temporal bone studies (e.g., Di Stadio et al., 2020; Fayad et al., 1991; Fayad & Linthicum, 2006; Heshmat et al., 2020; Kumar et al., 2022; Kusunoki et al., 2004; Linthicum & Fayad, 2009; Makary et al., 2011; Merchant et al., 2005; Nadol, 1990, 1997; Nadol et al., 1989; Rask-Andersen et al., 2010; Suzuka & Schuknecht, 1988; Ungar et al., 2018; Wu et al., 2019; Xing et al., 2012). These deteriorations start with damages in the myelin sheath and peripheral axon degeneration (e.g., Heshmat et al., 2020; Kumar et al., 2022; Nadol, 1990; Wu et al., 2019; Xing et al., 2012). Damaged spiral ganglion neurons (SGNs) with only central axons (i.e., unipolar SGNs) can survive decades after peripheral axon loss (e.g., Kusunoki et al., 2004; Linthicum & Fayad, 2009; Nadol, 1990; Rask-Andersen et al., 2010) and still be activated by electrical stimulation (e.g., Javel & Shepherd, 2000; Shepherd & Hardie, 2001; Shepherd & Javel, 1997; Sly et al., 2007; van den Honert & Stypulkowski, 1984). Eventually, the SGN soma and the central axon degenerate, which leads to the disappearance of the entire SGN (e.g., Fayad et al., 1991; Fayad & Linthicum, 2006; Linthicum & Fayad, 2009; Suzuka & Schuknecht, 1988; Ungar et al., 2018). The number and the distribution of surviving SGNs, the number of bipolar vs unipolar SGNs, as well as the degree of axonal degeneration and demyelination of remaining SGNs, vary substantially along the cochlea within and across CI patients (Fayad et al., 1991; Fayad & Linthicum, 2006; Linthicum & Fayad, 2009; Merchant et al., 2005; Nadol, 1997).

Deteriorations in anatomical structures of the CN reduce its discharge synchronization (i.e., neural synchrony). Specifically, both axonal dystrophy and demyelination alter many neural properties, such as membrane capacitance and resistance, nodal leakage resistance, as well as nodal sodium and potassium channel permeability (e.g., Tasaki, 1955; Waxman & Ritchie, 1993). These changes cause a reduction in the nodal current density, axonal spiking probability and propagation velocity, as well as an increase in temporal jitter, spike latency, and conduction vulnerability of individual CN fibers (e.g., Gonzalez-Gonzalez & Cazevieille, 2019; Heshmat et al., 2020; Kim et al., 2013; Tasaki, 1955). CN fibers with different degrees of axonal dystrophy and demyelination generate and conduct action potentials at different speeds, which reduces the synchronized discharge across the population of CN fibers (Kandel, 2002). Animals with more demyelination show greater reductions in neural synchrony in the CN (e.g., El-Badry et al., 2007). In electrical hearing, loss of the peripheral axon and altered membrane properties can also move the action potential initiation site distally to the SGN soma or central axon (e.g., Hartmann et al., 1984; Javel & Shepherd, 2000; van den Honert & Stypulkowski, 1984). Compared with responses initiated at peripheral axons, spikes initiated at central axons have less temporal dispersion or jitter (Javel & Shepherd, 2000). The difference in the action potential initiation site among CN fibers could further reduce discharge synchronization across CN fibers. Due to the lack of noninvasive tools to evaluate neural synchrony in the CN to electrical stimulation in CI users in the past, our knowledge in this area is primarily based on the results showing the variance in the first spike latency after stimulus onset (i.e., temporal jitter) of individual CN fibers measured using single fiber recordings in animal models (e.g., Hartmann et al., 1984; Parkins, 1989; Shepherd & Hardie, 2001; Shepherd & Javel, 1997; Sly et al., 2007; van den Honert & Stypulkowski, 1984). How well this knowledge applies to human CI users remains unknown due to the differences in anatomical/morphometric and biophysical properties of CN fibers, as well as durations and etiologies of deafness between human listeners and experimental animals (Skidmore et al., 2022). In addition, these results do not provide any information about discharge synchronization across electrically stimulated CN fibers or discharge synchronization of a group of CN fibers across repeated stimulations. To date, neural synchrony in the electrically stimulated CN in human listeners has not yet been evaluated and remains unknown. Its role in processing temporal cues and understanding speech in noise in CI users also remains unknown despite the rich literature showing its importance for these processes in acoustic hearing.

To address these critical knowledge gaps, we recently developed a noninvasive, *in vivo* method for assessing neural synchrony of a population of electrically stimulated CN fibers by quantifying the trial-to-trial phase coherence in the summated activity to electrical stimulation using electrophysiological measures of the electrically evoked compound action potential (eCAP). Using this new method, we studied the effect of peripheral neural synchrony on temporal resolution acuity by assessing the association between the degree of peripheral neural synchrony and within-channel gap detection threshold (GDT) measured using psychophysical procedures. The association between the degree of peripheral neural synchrony and Consonant-Nucleus-Consonant (CNC) word scores measured in quiet and in noise was also evaluated. These experiments were designed to test the hypothesis that peripheral neural synchrony is an important factor for temporal resolution acuity and speech perception outcomes in noise in post-lingually deafened adult CI users.

## MATERIALS AND METHODS

### Study Participants

This study included 24 (14 Female, 10 Male) post-lingually deafened adult CI users ranging in age from 36.8 to 84.0 years (mean: 63.7 years, SD: 12.8 years). All study participants were native speakers of American English and used a Cochlear™ Nucleus® device (Cochlear Ltd, Macquarie, NSW, Australia) with a full electrode insertion in the test ear, as confirmed based on post-operative, high-resolution computerized tomography scans. Participants A3, A5, and A12 were implanted bilaterally. For these three participants, each ear was tested separately. None of these participants has any functional acoustic hearing in either ear. eCAPs were measured in each of 27 ears tested in these 24 participants. Participant A16 was unable to participate in the speech perception evaluation. Participants A10, A16, A18 and A20 were not able to participate in psychophysical measures of GDT due to their limited availabilities. As a result, speech perception was evaluated for each of 26 ears tested in 23 participants. Psychophysical GDTs were measured at two CI electrodes in each of 23 ears tested in 20 participants. Detailed participant demographic information and the experiments that each participant completed are provided in Table 1. Written informed consent was obtained from all study participants at the time of data collection. The study was approved by the Biomedical Institutional Review Board (IRB) at The Ohio State University (IRB study #: 2017H0131).

**TABLE 1.**
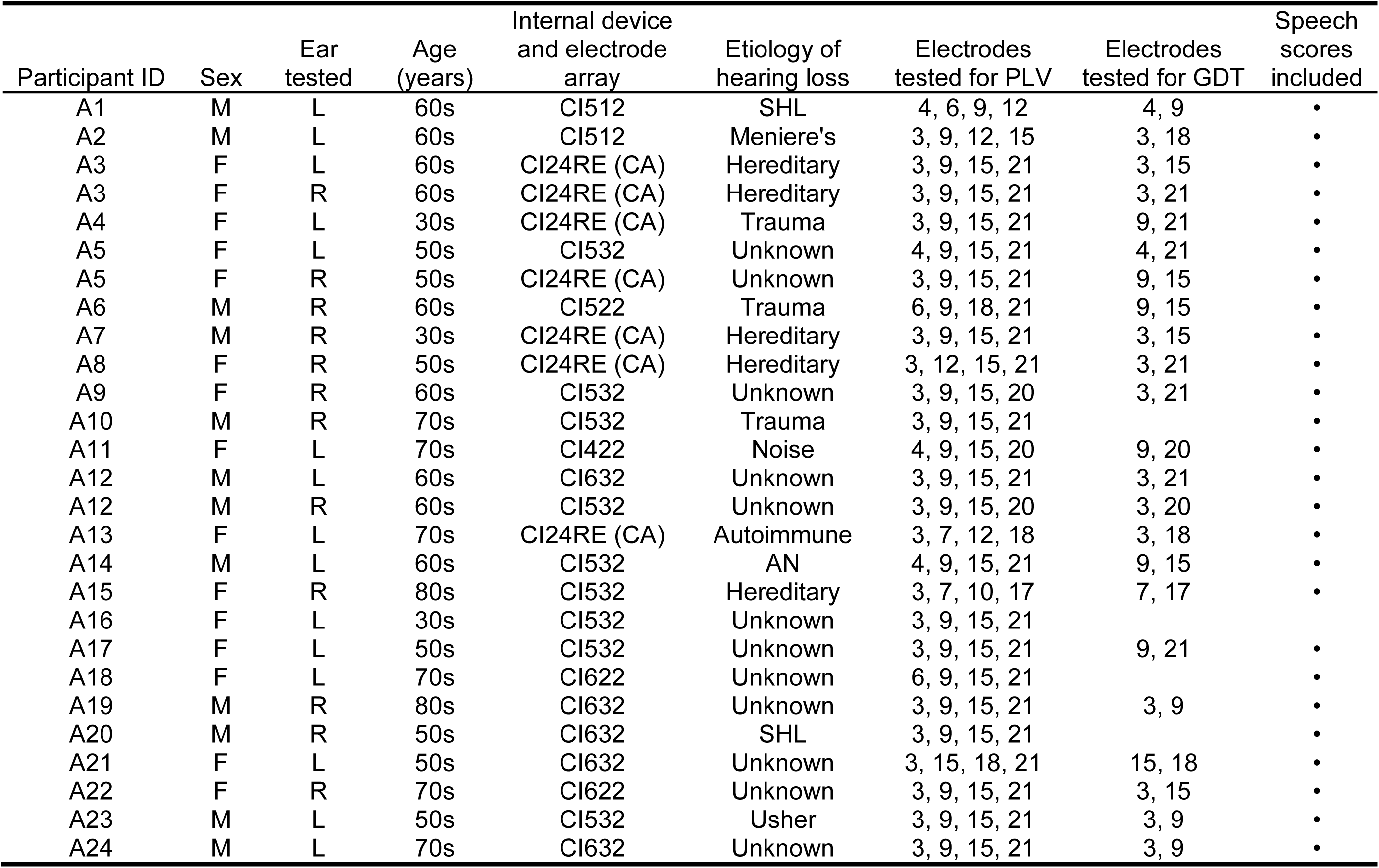
Demographic information of all study participants. CI24RE (CA), Freedom Contour Advance electrode array; SHL, sudden hearing loss; AN, acoustic neuroma

### Stimuli

For eCAP recording, the stimulus was a charge-balanced, cathodic-leading, biphasic pulse with an interphase gap of 7 µs and a pulse phase duration of 25 µs/phase. For measuring psychophysical GDT, the stimulus was a train of biphasic pulses with the same characteristics as those of the single-pulse stimulus that was presented for 500 ms at a stimulation rate of 900 pulses per second (pps) per channel. For both measures, the stimulus was delivered to individual CI electrodes in a monopolar-coupled stimulation mode via an N6 sound processor interfaced with a programming pod.

### Behavioral C Level Measures

The maximum comfortable level (i.e., the C level) for each type of stimulus was determined using an ascending procedure. In this procedure, study participants were instructed to use a visual loudness rating scale [scale of 1-10, where 1 is “barely audible” and 10 is “very uncomfortable”] to indicate when the sound reached the maximum comfort level (rating of 8). Stimulation was first presented at a relatively low level and gradually increased in steps of 5 clinical units (CUs) until a loudness rating of 7 was reached. Then, stimulation was increased in steps of 1-2 CUs until a rating of 8 (“maximal comfort”) was reached. The C level was measured for each type of stimulation delivered to each test CI electrode for each participant.

For the single-pulse stimulation used for eCAP recording, the stimulus was presented to individual CI electrodes using the “Stimulation Only” mode in the Advanced Neural Response Telemetry (NRT) function implemented in the Custom Sound EP (v. 6.0) commercial software (Cochlear Ltd, Macquarie, NSW, Australia) software. Due to the challenge of reliably rating loudness for an extremely brief single pulse, the C level was determined for a group of five pulses presented at 15 Hz. This is a standard clinical practice for determining the C levels during the programming process. For the pulse-train stimulation, the stimulus was presented to individual CI electrodes using a custom script prepared using Nucleus Interface Communicator Routine Library (NIC v. 4.3.1) (Cochlear Ltd, Macquarie, NSW, Australia).

### eCAP Measurements

The eCAP recordings were obtained using the NRT function implemented in the Custom Sound EP (v. 6.0) commercial software (Cochlear Ltd, Macquarie, NSW, Australia). The eCAP was measured at individual CI electrode locations using a two-pulse forward-masking-paradigm (Brown et al., 1990). In this paradigm, the masker and the probe pulse were presented to the test electrode at the participants’ C level and 10 CUs below the C level, respectively. The stimulation was presented 400 times using a probe rate of 15 Hz to minimize the potential effect of long-term adaptation on the eCAP (Clay & Brown, 2007). The number of trials was chosen to be 400 because preliminary analyses indicated that this number of trials could ensure accurate estimation of neural synchrony in the CN while maintaining a feasible recording time (Skidmore et al., 2023a). Results of one of our previous studies have demonstrated that using electrophysiological results measured at single CI electrode locations to correlate with auditory perception outcomes in CI users can lead to inaccurate, if not wrong, conclusions (He et al., 2023). In comparison, the averaged results across multiple testing electrode locations provide a better representation of overall neural function than the result measured at any individual CI electrode locations. Based on these results, four electrodes across the electrode array were tested for each participant to get an estimate of overall CN function while maintaining a feasible testing time. The default testing electrodes were electrodes 3, 9, 15, and 21. Alternate electrodes were tested in cases where there was an open- or short-circuit at the default electrode locations. The electrodes tested for each participant are listed in Table 1. Other parameters used to record the eCAP included a recording electrode located two electrodes away apically from the stimulating electrode except for electrode 21 which was recorded at electrode 19, a 122-µs recording delay, an amplifier gain of 50 dB, and a sampling rate of 20,492 Hz.

### Measure of Neural Synchrony

Peripheral neural synchrony at individual CI electrode locations was evaluated based on 400 individual sweeps (trials) of the eCAP. Neural synchrony was quantified using an index named the phase locking value (PLV) which is a measure of trial-to-trial phase coherence among the 400 eCAP sweeps. The PLV is a unitless quantity that ranges from 0 to 1. It quantifies the degree of synchrony in neural responses generated by target CN fibers across multiple stimulation/trials. The PLV is influenced by temporal jitter in spike firing of individual CN fibers, discharge synchronization across the population of activated CN fibers, and discharge synchronization of a group of activated CN fibers across multiple stimulations. A PLV of 0 means that the distribution of phase across trials is uniform (i.e., the responses across trials are uncorrelated). The PLV is 1 if phases across trials are perfectly correlated. As a result, larger PLVs indicate better/stronger neural synchrony in the CN. Mathematically, the PLV is the length of the vector formed by averaging the complex phase angles of each trial at individual frequencies obtained via time-frequency decomposition. Specifically, the PLV is calculated at a specific frequency and time window (i.e., frame) as

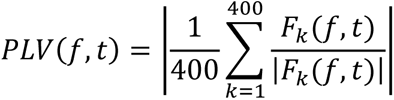

where *F*_*k*_(*f*, *t*) is the spectral estimate (i.e., complex number representing the amplitude and phase of a sinusoid obtained from the short-time Fourier transform) of trial *k* at frequency *f* for the time window *t*. For this study, the time-frequency decomposition was performed at six linearly spaced frequencies (788.2, 1576.3, 2364.4, 3152.6, 3941.0 and 4729.2 Hz) with Hanning Fast Fourier Transform tapers, a pad-ratio of 2 and a frame size of 26 samples (1268.8 µs) using the newtimef function (v. 2022.1) included in the MATLAB plugin EEGlab (Delorme & Makeig, 2004). For each CI electrode tested in each participant, a single PLV was obtained by averaging PLVs calculated at six frequencies for six partially overlapped frames with an onset-to-onset interval of 48.8 μs between two adjacent frames within a time window of 1561.6 μs. The use of six partially overlapped frames within the time window of interest allows for higher temporal resolution of the PLV, with PLV values in the early frames capturing the degree of synchrony in the low-latency spikes and values in the later frames being dependent on synchrony in the longer-latency spikes.

The parameters used in PLV calculation were selected based on morphological characteristics of the eCAP measured in human CI users and the sampling rate offered by CI manufacturers for eCAP recording. Specifically, the eCAP recorded in human CI users consists of one negative peak (N1) within a time window of 0.2 – 0.4 ms after stimulus onset followed by a positive peak (P2) occurring around 0.6 – 0.8 ms (for a review, see He et al., 2017). As a result, the longest inter-peak latency of the eCAP in human listeners is 600 µs. Using this duration as the half width of the sinusoid included in Fast Fourier Transform analysis and the sampling rate of 20,492 Hz offered by Cochlear™ Nucleus® device for measuring the eCAP, the frame size was determined to be 1268.8 µs in time which included 26 samples (1268.8/48.8 = 26). As a result, six frames with an onset-to-onset interval of 48.8 μs between two adjacent frames cover the entire recording window [(1561.6-1268.8)/48.8 = 6]. This frame size in time also determines the lowest frequency (1/1268.8 µs = 788.1 Hz) used in the PLV calculation. The difference in the peak-to-baseline amplitude between the N1 and the P2 peak of the eCAP and the difference in their widths indicate a complex spectrum instead of a single fundamental frequency. A Fast Fourier Transform analysis was conducted to determine the frequency components of the averaged eCAPs over 400 sweeps measured in this study. Results showed that the harmonic frequency with one quarter of the amplitude measured at the fundamental frequency was 4482.6 Hz. As a result, the highest frequency used in PLV calculation was determined to be 4729.2 based on a spectral resolution of 788.1 Hz determined by the frame size in time. This frequency range (i.e., 788.1 ‒ 4729.2 Hz) is higher than that used in Harris et al. (2018, 2021). This difference is caused by morphological differences between the eCAP and the compound action potential evoked by acoustic clicks.

Figures 1 and 2 use example data recorded at four CI electrode locations in participant A14 and in the right ear of participant A3 to illustrate this method. These examples were chosen because they included the lowest and the highest PLV measured in this study. Electrode number, the resulting PLV and the amplitude of the eCAP averaged over 400 sweeps/trials are indicated in each panel. It should be noted that these eCAPs were measured at the C level measured for each tested CI electrode in each participant. Therefore, the variation in stimulation levels used to measure the eCAP and the difference in corresponding eCAP amplitudes across different CI electrodes within each participant are due to participant-related factors instead of measurement bias.

**Figure 1.**
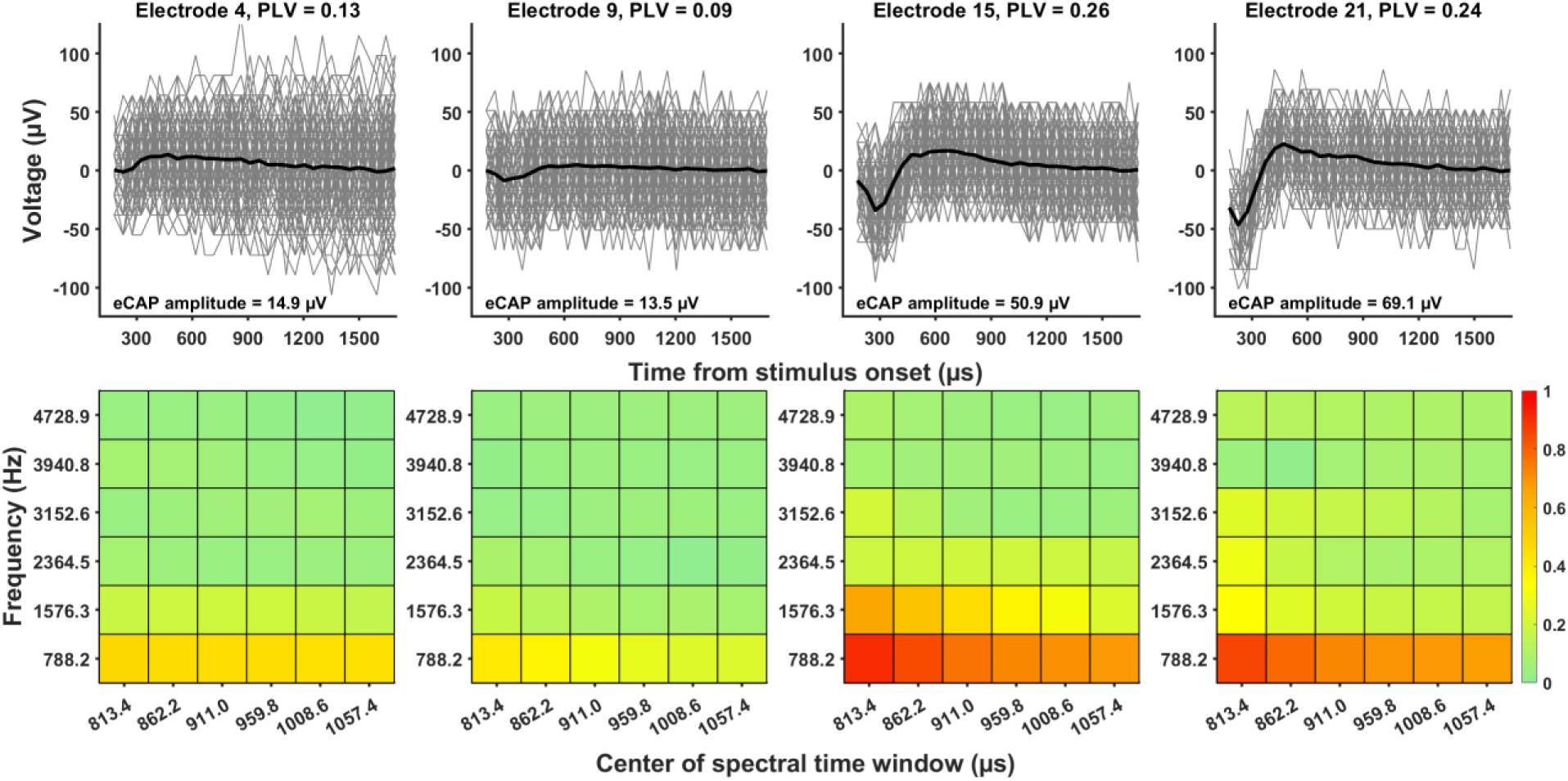
Representative data from participant A14 demonstrating the method for estimating neural synchrony at the level of the cochlear nerve at individual electrode locations in cochlear implant users. Top panels: Recordings of electrically evoked compound action potentials (eCAPs) for individual trials (gray lines) with the across-trial average (black line). The amplitude of the across-trial average is also provided. Bottom panels: Heat maps indicating the phase-locking value (PLV) as a function of time and frequency.

**Figure 2.**
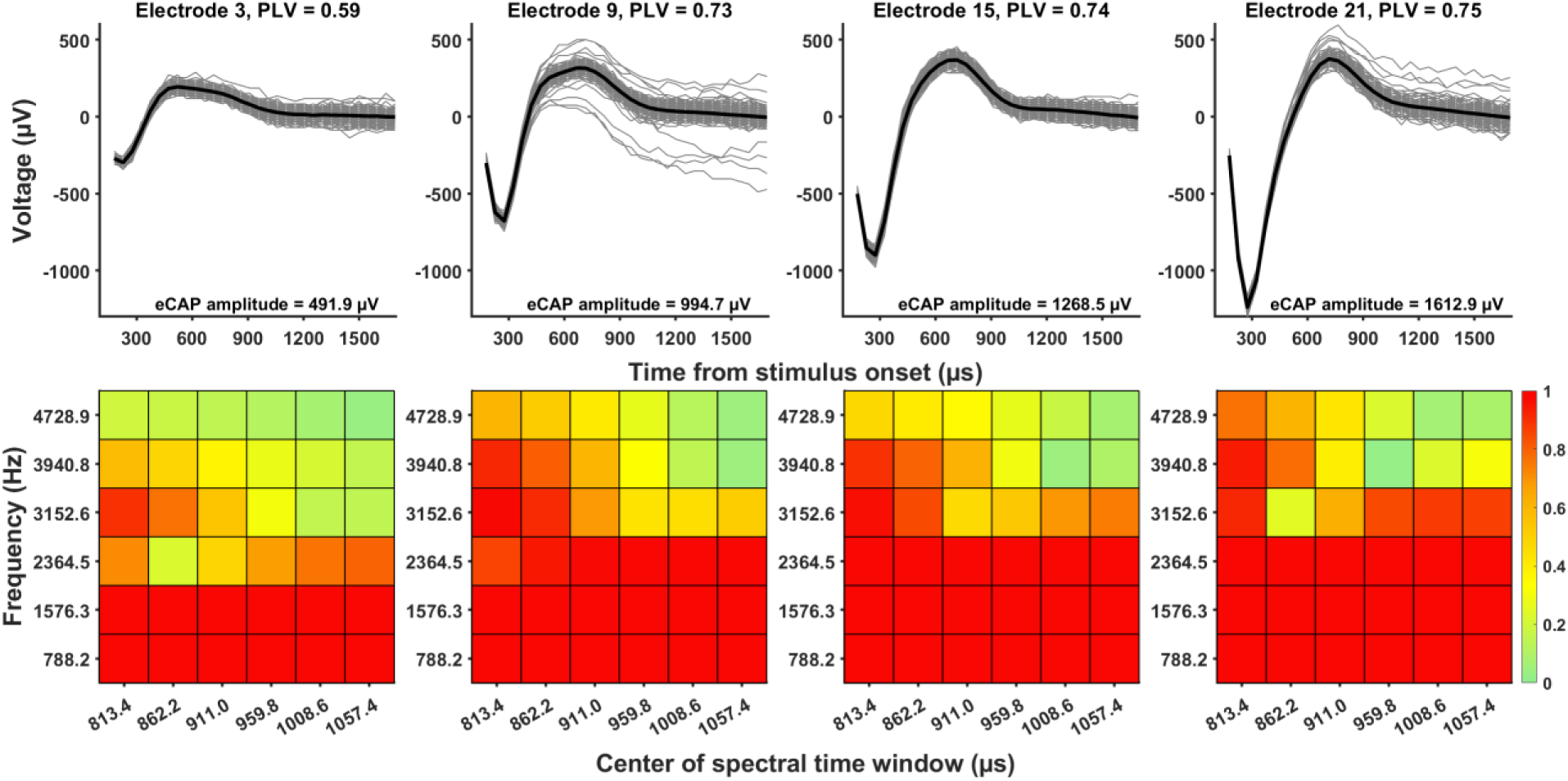
Representative data recorded in the right ear of participant A3 demonstrating the method for estimating neural synchrony at the level of the cochlear nerve at individual electrode locations in cochlear implant users. Top panels: Recordings of electrically evoked compound action potentials (eCAPs) for individual trials (gray lines) with the across-trial average (black line). The amplitude of the across-trial average is also provided. Bottom panels: Heat maps indicating the phase-locking value (PLV) as a function of time and frequency.

### Psychophysical Measures of Gap Detection Threshold

Within-channel GDTs were measured at two electrode locations with different PLVs in each of 23 ears tested in 20 participants (see Table 1 for the electrodes tested in each ear). Pulse trains with and without temporal gaps were presented in a three-alternative, forced-choice paradigm that incorporated a three-down, one-up adaptative strategy to estimate 79.4% correct on the psychometric function (Levitt, 1971). Individual trials consisted of three consecutive 500-ms listening intervals separated in time by 500 -ms silent intervals. The stimulus presented in two of the three listening intervals was a 500-ms pulse train without any interruption. The stimulus presented in the remaining listening interval, chosen at random, included a temporal gap centered at 250 ms of stimulation. The participant was asked to determine which of the three listening intervals included two sounds. Feedback on correct/incorrect choices was not provided to participants. The gap duration began at 64 ms and was shortened/lengthened based on the correctness of the participants’ choice. The initial step size of the change in gap duration was 32 ms. This step changed by a factor of two after three consecutive correct responses or one incorrect response. The minimum and maximum gap durations permitted were 1 ms and 256 ms, respectively. The GDT was calculated as the average across two trials in which the mean gap duration over the last four (of twelve) reversals was calculated.

### Speech Measures

Speech perception performance was evaluated separately for each implanted ear using Consonant-Nucleus-Consonant (CNC) word lists (Peterson & Lehiste, 1962) presented in quiet and in two noise conditions. All auditory stimuli were presented in a sound-proof booth via a speaker placed one meter in front of the subject at zero degrees azimuth. The target stimulus was always presented at 60 dB sound pressure level (SPL). For the noise conditions, speech-shaped noise was presented concurrently with the target stimulus at 50 dB SPL and 55 dB SPL to create signal-to-noise ratios of +10 dB and +5 dB, respectively.

### Statistical Analyses

Descriptive statistics of PLVs measured at different electrode locations, GDTs and CNC word scores measured in different testing conditions and the degree of noise effect on CNC word scores which was quantified as the difference in CNC word scores measured in quiet and in noise were calculated, including the overall mean and standard deviation. Effects of electrode location and stimulation level on the PLV were assessed using a Linear Mixed effect Model (LMM) with electrode location and stimulation level as fixed effects. The effect of the PLV on GDT was evaluated using a LMM with the PLV, the stimulation level used to measure the GDT and the stimulation used to measure the PLV as fixed effects. All LMMs used a correlated regression model with an unstructured correlation matrix to account for repeated observations per participant. Estimations were obtained using restricted maximum likelihood with Satterthwaite degrees of freedom. The Tukey’s Honest Significant Difference (Tukey’s HSD) method was used to adjust for multiple comparisons. The difference in GDT between results measured at the two electrodes with different PLVs or stimulation levels were assessed using paired sample t-tests. One-tailed Pearson product-moment correlation tests with Bonferroni correction for multiple comparisons were used to assess the association of the PLV with CNC word scores measured in different conditions (α = 0.017), as well as with the change in CNC word score with competing background noise (i.e., CNC word score measured in quiet - CNC word score measured in noise, α = 0.025). Using electrophysiological results measured at single CI electrode locations to correlate with auditory perception outcomes in CI users can lead to inaccurate conclusions (He et al., 2023). Therefore, for these correlation analyses, PLVs measured at all electrode locations were averaged together for each participant/ear to obtain an estimation of the overall peripheral neural synchrony within the cochlea and to minimize electrode-location related bias in study results. One-tailed Pearson product-moment correlation tests with Bonferroni correction for multiple comparisons were also used to assess the association between CNC word scores measured in quiet and the degree of noise effect on CNC word scores. The strength of correlation was determined based on values of the Pearson product-moment correlation coefficient (r). Specifically, weak, moderate, and strong correlations were defined as *r* values between 0 and 0.3 (0 and −0.3), between 0.3 and 0.7 (−0.3 and −0.7), and between 0.7 and 1.0 (−0.7 and −1.0), respectively.

All statistical analyses for this study were performed using R software (v. 4.3.0) (R Core Team, 2021). All statistical models were fitted using the nlme package (Pinheiro et al., 2023) and pairwise comparisons were evaluated using the emmeans package (Lenth, 2023).

## RESULTS

### Neural Synchrony Along the Cochlea

PLVs measured in this study ranged from 0.09 to 0.76 (mean: 0.55, SD: 0.22) across all electrodes tested. The means and standard deviations of PLVs measured at each of the four electrode locations are shown in Figure 3. The standard deviations at all four electrode locations demonstrate variations in the PLV in CI users. In addition, a trend for the mean PLV to be different across electrode locations is observed. PLVs measured at apical electrode locations (i.e., electrodes 15 and 21) appear to be larger than those measured at more basal electrode locations (i.e., electrodes 3 and 9). The results of the LMM showing a significant effect of electrode location on the PLV 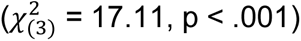 after controlling for the significant effect of stimulation level on the PLV (t_(101)_ = 3.90, p < .001). The results of pairwise comparisons showed that PLVs measured at electrode 15 were significantly larger than those measured at electrode 3 (t_(26.9)_ = −3.46, p = .009) and electrode 9 (t_(27.8)_ = −3.62, p = .006). The differences in the PLV measured between other electrode pairs did not reach a statistical significance. Detailed results of pairwise comparisons are listed in Table 2.

**Figure 3.**
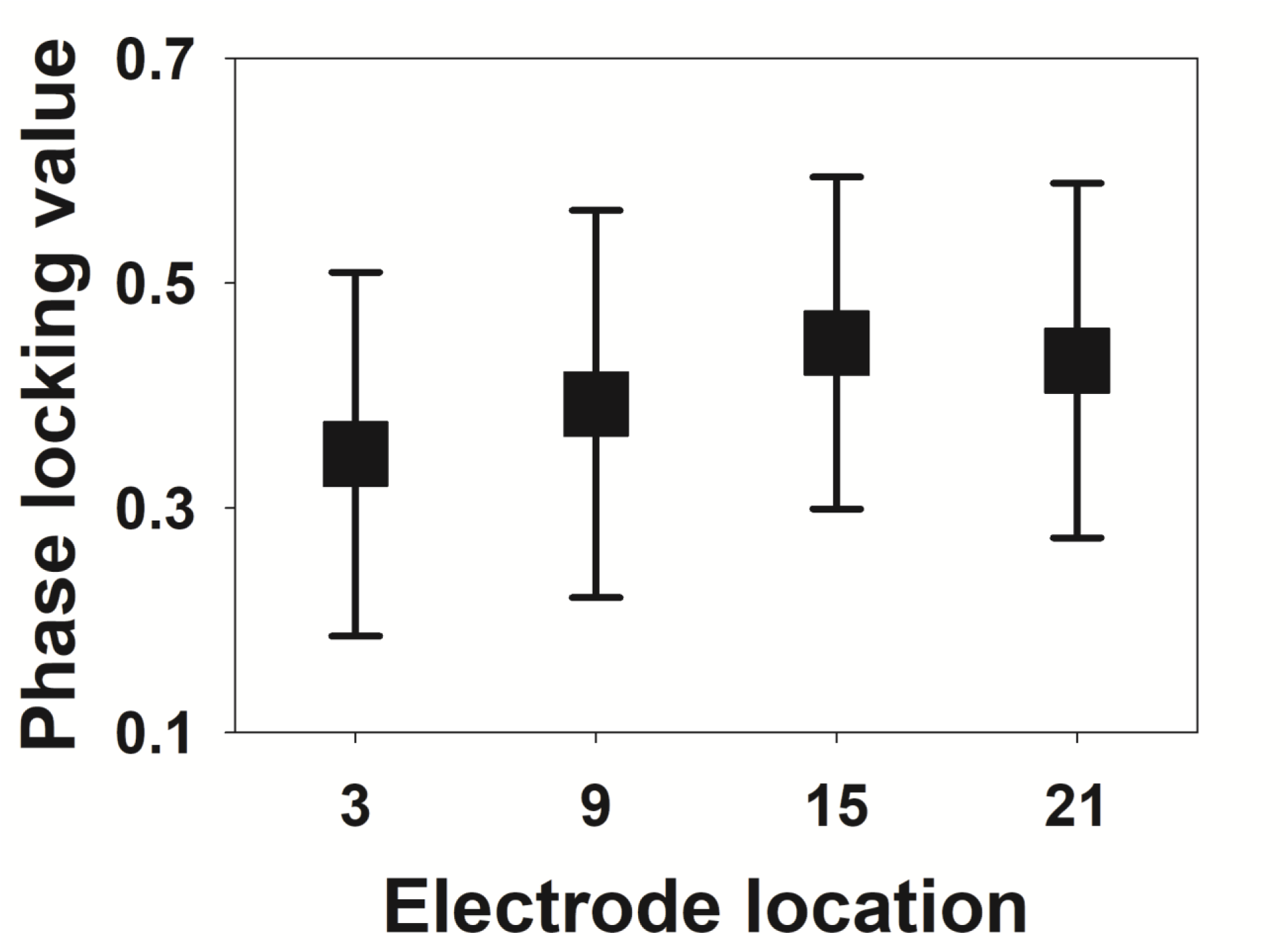
The means and standard deviations of phase-locking values measured at four electrode locations in 24 adult cochlear implant users (27 ears).

**Table 2.** Results of pairwise comparisons for comparing phase locking values measured at different electrode locations.

| Electrode Pair | Estimate | Standard Error | Degree of Freedom | T Ratio | *p* Value |
| --- | --- | --- | --- | --- | --- |
| E3 vs. E9 | -0.022 | 0.021 | 30.2 | -1.025 | .736 |
| E3 vs. E15 | -0.080 | 0.023 | 26.9 | -3.463 | .009 |
| E3 vs. E21 | -0.087 | 0.031 | 30.9 | -2.865 | .035 |
| E9 vs. E15 | -0.059 | 0.016 | 27.8 | -3.622 | .006 |
| E9 vs. E21 | -0.066 | 0.028 | 33 | -2.575 | .067 |
| E15 vs. E21 | 0.007 | 0.020 | 30.9 | -0.352 | .984 |

### Neural Synchrony and Temporal Acuity

Figure 4 shows psychophysical GDTs measured at two electrode locations per test ear in 20 participants (23 ears) as a function of the PLV. For the two CI electrode locations tested in each of 19 ears in 18 participants, smaller GDTs were always measured at the electrode locations with higher PLVs. Results measured in A8, A24 and the right ear of A5 showed an opposite pattern. In the left ear of A12, GDTs measured at the two CI electrode locations with different PLVs were the same (5.75 ms). The result of a paired-samples t-test showed that GDTs measured at the electrode locations with larger PLVs were statistically significantly smaller than those measured at electrode locations with smaller PLVs (t_(22)_ = 2.25, p = .035). The results of the LMM showed a significant effect of the PLV on GDT (t_(42)_=-3.51, p = .001) after controlling for the stimulation level of GDT (t(42) = −2.53, p = .015) and the stimulation level of the PLV (t_(42)_ =3.75, p < .001), with larger PLVs leading to smaller GDTs.

**Figure 4.**
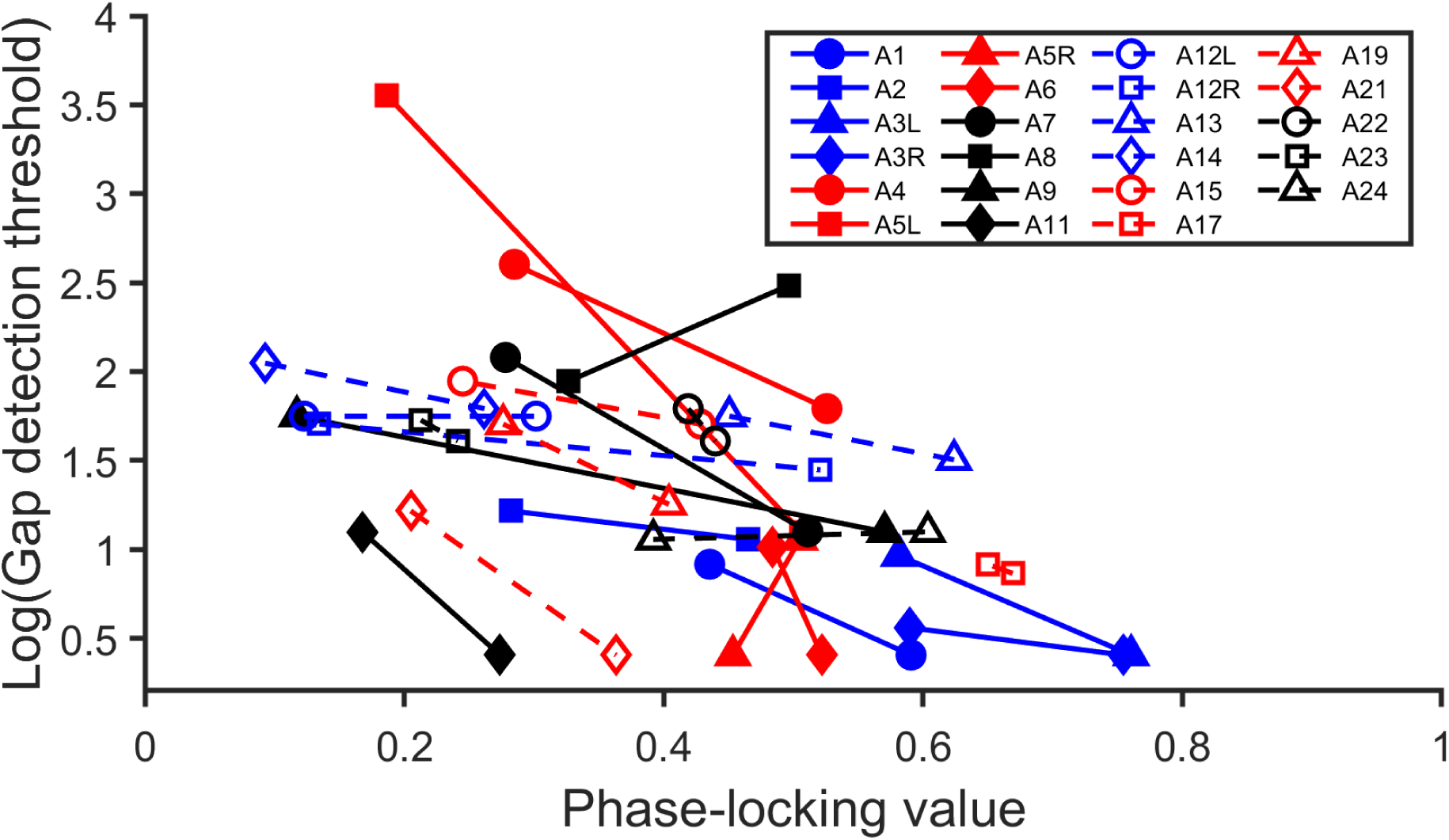
Phase-locking values and psychophysical gap detection thresholds measured at two electrode locations in each of 23 implanted ears of 20 participants. Lines connect the data measured at the two electrode locations tested in the same ear.

Figure 5 shows psychophysical GDT results as a function of stimulation levels used to measure these GDTs. Smaller GDTs were measured at higher stimulation levels for the two CI electrode locations tested in each of 10 ears tested in 9 participants. Results measured in 11 ears tested in 11 participants showed an opposite relation between these two parameters. For A2, the stimulation levels used to measure GDTs at electrodes 3 and 18 were the same (196 CU). For A12, using different stimulation levels at electrodes 3 and 21 resulted in the same GDT (i.e., 5.75 ms). Overall, despite a significant stimulation level effect on GDT at a group level as shown in the LMM results, these data do not demonstrate a consistent association between stimulation level and GDT across participants, which differs from those shown in Figure 4. Consistent with these observations, the result of a paired-samples t-test showed that GDTs measured at the electrode locations with higher stimulation levels were not significantly different from those measured at the electrode locations with lower stimulation levels (t_(22)_ = −0.53, p = .599).

**Figure 5.**
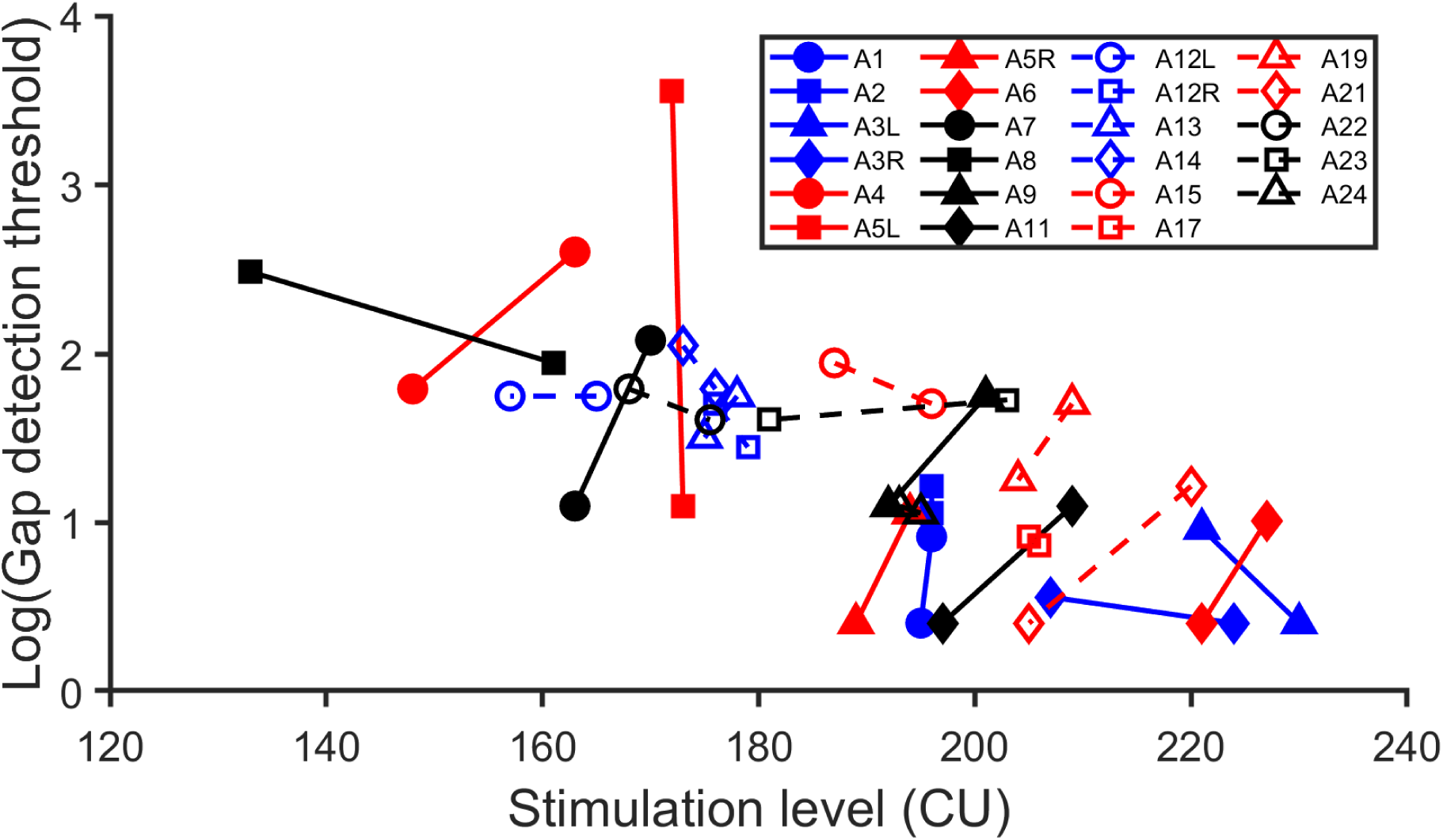
Stimulation levels and psychophysical gap detection thresholds measured at two electrode locations in each of 23 implanted ears of 20 participants. Lines connect the data measured at the two electrode locations tested in the same ear.

### Neural Synchrony and Speech Perception

Figure 6 shows CNC word scores measured in quiet and in two noise conditions as a function of the PLV for 23 participants (26 ears). There was substantial variability in CNC word scores measured in quiet (range: 40.0 – 96.0%, mean: 73.1%, SD: 13.3%) and in noise (+10 dB SNR: range: 22.0 – 84.0%, mean: 58.1%, SD: 15.8%; +5 dB SNR: range: 8.0 – 82.0%, mean: 45.6%, SD: 15.6%). There was no obvious relation between CNC word score and the PLV for each of the testing conditions. This observation was confirmed by the result of a Pearson product-moment correlation test with Bonferroni correction for multiple testing (Quiet: r = −0.18, p = .193; +10 dB SNR: r = −0.08, p = .353; +5 dB SNR: r = 0.15, p = .238).

**Figure 6.**
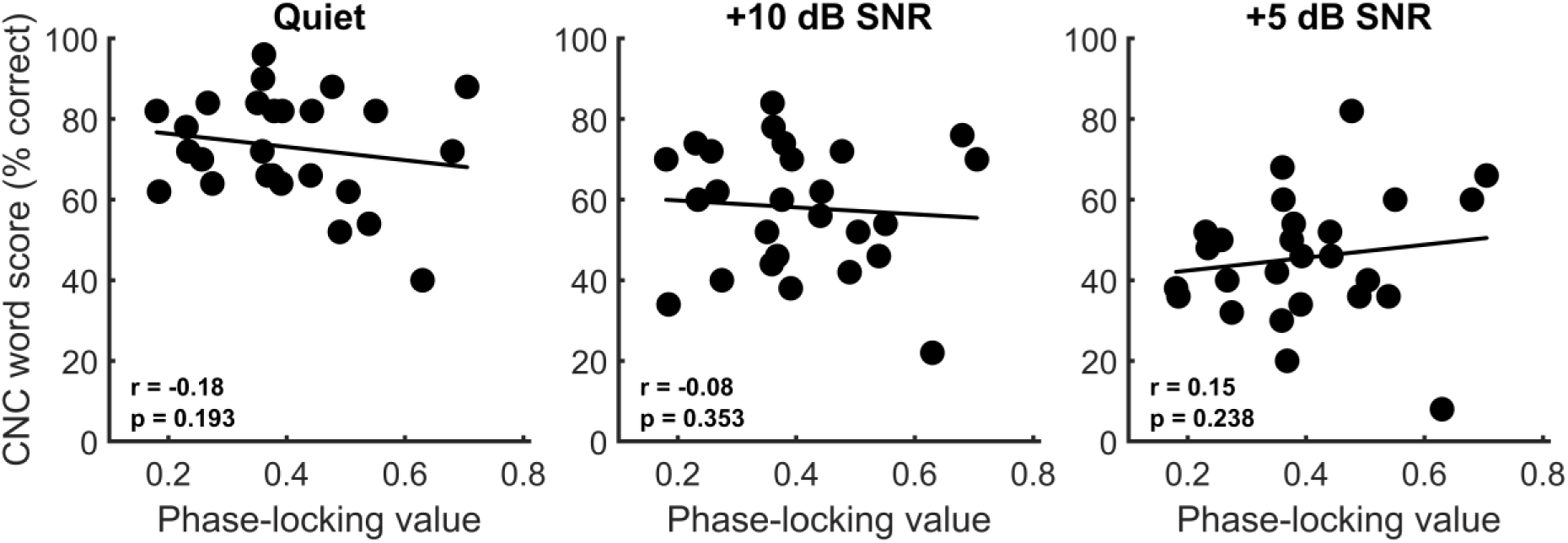
Consonant-Nucleus-Consonant (CNC) word scores measured in quiet and in two noise conditions as a function of the phase-locking value averaged across electrode locations for 23 adult cochlear implant users (26 ears). The best fit line across all 26 data points is illustrated with a solid line. The results from Pearson’s correlation analysis are also provided in each panel.

A careful inspection of study results showed that the amount of change in CNC word scores with the presence of noise also varied among CI users (+10 dB SNR: range: −32 ‒ 4.0%, mean: −15.0%, SD: −9.5%; +5 dB SNR: range: −46.0 – −6.0%, mean: −27.4%, SD: −10.9%). For individual participants, the amount of change could not be predicted based on their scores measured in quiet. For example, the CNC scores measured in quiet in the right ear of participants A5 (A5R) and A19 (A19R) were 88% and 84%, respectively. While A19R showed a 44% decrease in CNC word score when a noise at +5 dB SNR was added, A5R only had a 6% decease. Similarly, a CNC score of 72% measured in quiet was obtained for A3L and A15R. While A3L showed a 12% decrease in CNC word score, A15R had a 42% decrease when a noise at +5 dB SNR was added. Finally, both A7R and A17L showed a 32% decrease in CNC word scores when a noise at +5 dB SNR was added despite a 56% difference in CNC word scores measured in quiet between these two cases (scores measured in quiet: A7R: 96%, A17L: 40%). These observations were confirmed by the results of Pearson product-moment correlation tests with Bonferroni correction for multiple testing showing the nonsignificant correlation between CNC word score measured in quiet and the change in CNC word score when noise was added (+10 dB SNR: r = −0.07, p = .365; +5 dB SNR: r = −0.18, p = .192). Due to these variations in the amount of change in CNC word scores with the presence of noise, participants with similar scores measured in quiet could show largely different scores measured in noise and *vice versa*. Overall, these results suggested individual variations in susceptibility to background noise among CI users, which could not be fully captured by their scores measured in noise.

To determine whether neural synchrony in the CN was a potential contributing factor to individual variations in noise susceptibility, we evaluated the relation between the PLV and the degree of noise effect on CNC word scores which was quantified as the amount of change in CNC word scores with the presence of noise. Figure 7 shows the change in CNC word scores plotted as a function of the PLV for both noise conditions. There was no obvious relation between the noise effect on CNC word score and the PLV for the +10 dB SNR noise condition, which was confirmed by the result of a Pearson product-moment correlation test (r = −0.12, p = .281).The result of a Pearson product-moment correlation test with Bonferroni correction for multiple testing showed a moderate, negative correlation between the PLV and the degree of detrimental effect of background noise on CNC word score for the results measured at a SNR of +5 dB (r = −0.42, p = .016), with larger PLVs associated with smaller negative effects of background noise. This correlation is statistically significant after adjusting for multiple comparisons.

**Figure 7.**
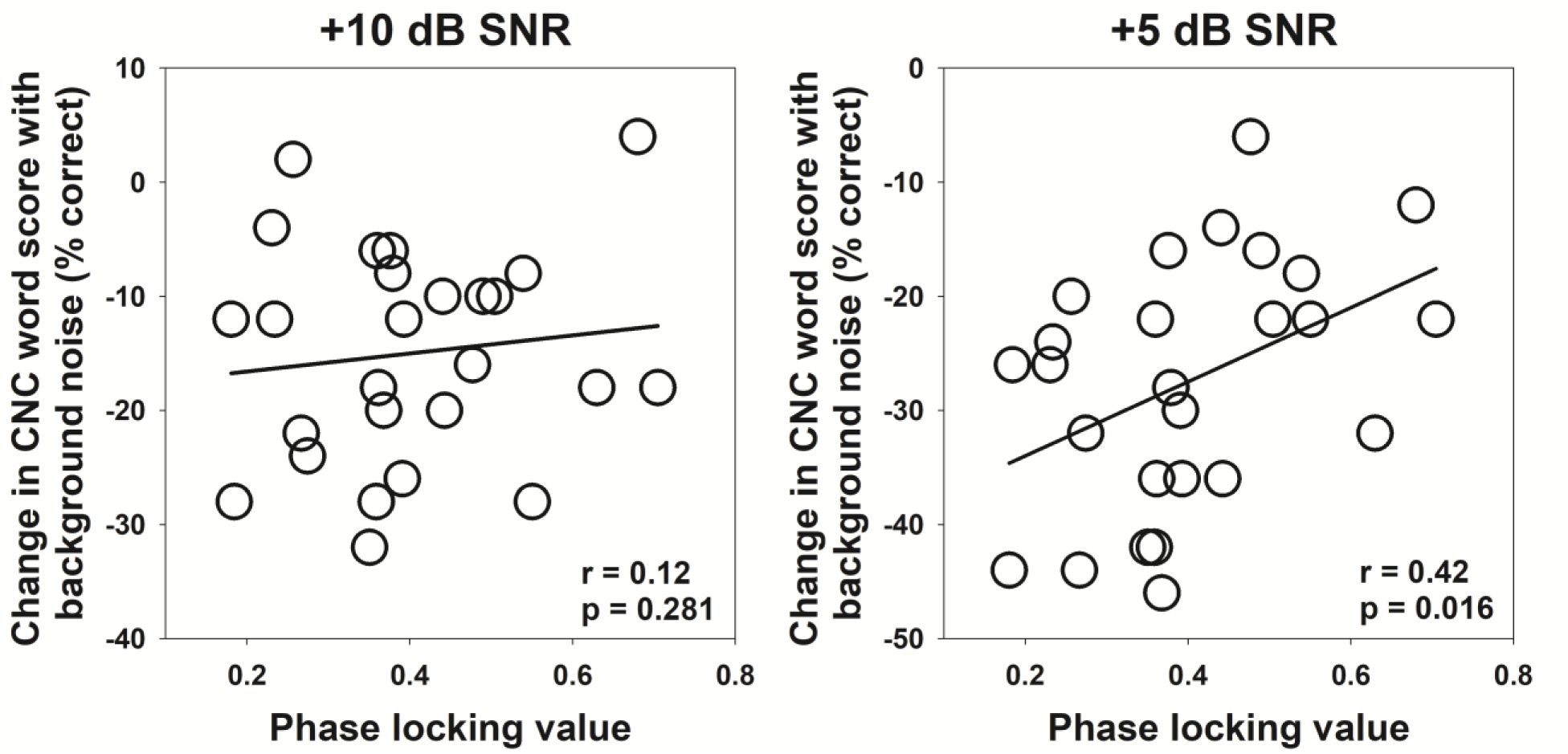
The change in Consonant-Nucleus-Consonant (CNC) word scores with the addition of background noise as a function of the phase-locking value averaged across electrode locations. The best fit line across all 26 data points is illustrated with a solid line. The results from Pearson’s correlation analysis are also provided in each panel.

### Periomodiolar vs. Lateral Wall Electrode Arrays

In this study, 16 ears of 15 participants were implanted with a periomodiolar electrode array and all other testing ears were implanted with a lateral wall electrode array. Even though it was not included in our original study design or the primary interest of this study, we conducted additional data analyses to determine whether the associations between the PLV, GDT and CNC word results differ between these two electrode arrays. Specifically, the effect of the PLV on GDT was evaluated using a LMM with the PLV, the stimulation level used to measure the PLV, the stimulation level used to measure GDT and electrode array type as fixed effects, participant as a random effect and an interaction between the PLV and electrode array type. The result showed a significant interaction (t_(73)_ = 2.55, p = .013), which suggests that the effect of the PLV on GDT differs between these two electrode arrays. The estimated slopes of linear regression functions modeling the relation between GDT and the PLV were significantly different from zero for both electrode arrays (lateral wall array: slope = −0.021, p < .001; periomodiolar array: slope = −0.006, p = .018). The difference in the estimated slopes between lateral wall and periomodiolar arrays was statistically significant (t_(33.7)_ = −2.55, p = .015), which evidences a stronger effect of the PLV on GDT for lateral wall electrode array than for periomodiolar electrode array.

The relation between the PLV and CNC word results by electrode array type was evaluated using linear regression analyses. The outcome variable was the PLV with linear predictors of CNC word results and electrode array type, and the interaction between electrode array type and CNC scores/changes in CNC scores with noise. The interaction term included in the linear regression was used to determine if the relation between CNC results and the PLV differed by electrode array type. Evaluations were conducted with a total of five regression models with each model built for each CNC result. Overall, none of these models showed a significant interaction between electrode array type and CNC results (p > .05). Therefore, there is no sufficient evidence to suggest that the relation between the PLV and CNC word results differs by electrode array type.

## DISCUSSION

This paper reports a newly developed method for quantifying neural synchrony in the electrically stimulated CN in human CI users. Using this newly developed method/tool, we evaluated the effects of peripheral neural synchrony on temporal resolution acuity and speech perception outcomes in human CI users. Results of this study showed variations in the degree of peripheral neural synchrony among CI users and demonstrated the important role that peripheral neural synchrony played in determining temporal resolution acuity in post-lingually deafened adult CI users. Our results also demonstrated a lack of association between the PLV and CNC word scores measured in quiet or in noise, as well as between the PLV and the amount of change in CNC word scores when a competing background noise at a SNR of +10 dB was added. However, there was a statistically significant negative correlation between the PLV and the degree of negative effect of background noise at a SNR of +5 dB. Overall, these results support our study hypothesis.

### Peripheral Neural Synchrony in Electrical Hearing

PLVs measured in CI users in this study ranged from 0.09 to 0.76, which is much higher than those measured in listeners with acoustic hearing (Harris et al., 2021). These results are consistent with the literature in animal models showing lower temporal jitters (i.e., higher discharge synchronization) of neural responses evoked by electrical stimulation than those evoked by acoustic stimulation (e.g., van den Honert & Stypulkowski, 1984). As a result, morphological characteristics of the eCAP are different from those of the compound action potential evoked by acoustic stimulation, which lead to the differences in frequency range, frame size and the number of linearly spaced frequencies used in Fast Fourier Transform analysis between this study and Harris et al. (2021). In addition, the recording window used in Harris et al. (2021) for PLV calculation is much longer than that used in this study (i.e., 10 ms vs 1561.6 μs). As a result, the number of frames used in these two studies are also different. Another important difference is how the final, single PLV is defined. In Harris et al. (2021), it was defined as the peak PLV across a 2-ms window around the N1 peak of the CAP. In our study, it was defined as the averaged PLV across six partially overlapped frames to better capture the degree of synchrony in spikes with varied latencies. All these factors could contribute to the different PLV ranges observed in these two studies. Note that our use of the mean PLV is more similar to the approach use in an earlier study by Harris and colleagues, in which they calculated a single PLV value by taking the *median* value across time and frequency (Harris et al., 2018).

Results of previous histological and functional studies have shown unpredictable patterns of CN health within and among typical CI users with heterogeneous etiologies (e.g., DeVries et al., 2016; Nadol et al., 2012; Sagers et al., 2017; Schvartz-Leyzac & Pfingst, 2016). Therefore, variations in peripheral neural synchrony would be expected across CI users and across electrode locations within individual patients, which is consistent with the wide ranges of PLVs being measured at different electrode locations across the cochlea in this study. Neural structures at the apex tend to be healthier than those located at more basal region of the cochlea in typical listeners with sensorineural hearing loss (Zimmermann et al., 1995), which could contribute to the higher PLVs measured at more apical locations in this study.

### Peripheral Neural Synchrony and Auditory Perception Outcomes

GDTs measured in this study are consistent with those reported in other studies that used similar testing paradigms and conditions (e.g., Busby & Clark, 1999; Garadat & Pfingst, 2011; Shader et al., 2020). Despite variations in GDTs and PLVs measured among participants, larger GDTs were observed at the electrode locations with smaller PLVs in all except for four implanted ears tested in this study. These data demonstrated that poor peripheral neural synchrony led to declined temporal resolution acuity in CI users, which is consistent with those measured in acoustic hearing (Michalewski et al., 2005; Zeng et al., 2005; Zeng et al., 1999). It should be noted that stimulation rate affects peripheral neural synchrony in electrical hearing, with higher stimulation rates resulting in lower/worse neural synchrony (Rubinstein et al., 1999). As a result, the degree of peripheral neural synchrony induced by the pulse train stimulation used to measure GDT is expected to be much lower than that quantified by the PLV in this study. Nevertheless, the results of both measures are affected by dyssynchronous neural firing, which explains to the strong relation between these two measures.

Our results showed a lack of correlation between peripheral neural synchrony and CNC word scores. These results are not consistent with the significant correlation between peripheral neural synchrony and CNC word scores measured in quiet reported by Dong et al. (2023). However, it should be noted that peripheral neural synchrony was not directly assessed in the experimental design of Dong et al. (2023). Instead, it was estimated using computational modeling techniques by deconvolving intraoperative recordings of the eCAP with an estimated human unitary response to obtain the distribution of firing latencies summed across CN fibers. As acknowledged by the authors, the estimation of neural synchrony using this modeling approach is highly dependent on the shape of the assumed unitary response from CN fibers, which has not been assessed/validated directly in humans (Dong et al., 2020; Dong et al., 2023). It is possible that degeneration of CN fibers leads to changes in the effective unitary response functions for each fiber, such that their simulated results did not fully reflect the actual peripheral neural synchrony in CI users. This methodological difference could contribute to the discrepancy between the results of Dong et al. (2023) and the present study. Speech perception outcomes in noise were not evaluated by Dong et al. (2023).

One finding of this study is the moderate correlation between the PLV and the detrimental effect of background noise on speech perception outcomes measured for the +5 dB SNR noise condition. This finding suggested that the degree of neural synchrony, as quantified using the PLV, accounted for approximately 18% of the negative effect of competing background noise presented at a SNR of +5 dB on CNC word scores. This statistically significant result could also be clinically meaningful given the fact that combining multiple factors could only explain less than 40% of variance in speech perception outcomes in CI users (Blamey et al., 1996; Blamey et al., 2013; Holden et al., 2013; James et al., 2019; Lazard et al., 2012). This association was not observed for speech perception outcomes measured at +10 dB SNR. Overall, these results indicated that peripheral neural synchrony could be an important factor determining the degree of the noise effect on speech perception outcomes in CI users in the case of mixed speech and masker signals presented to the same ear. The importance of peripheral neural synchrony to speech perception seems to increase with elevated background noise, which is consistent with our previous results showing a stronger impact of CN function on speech perception outcomes in more challenging listening conditions (Skidmore et al., 2023b).

### Methodological Considerations

There are several methodological factors that need to be considered when applying this method in future studies. First, using different parameters in mathematical calculation will result in different PLV results. To determine whether this is a crucial factor for this newly developed method, we used three additional sets of parameters to calculate the PLV and assessed their associations with GDTs, CNC word scores measured in different conditions and the noise effect on CNC word scores using the same statistical analysis methods as those reported in this study. Overall, PLVs calculated using all four sets of parameters are strongly correlated with each other (Pearson correlation coefficients: 0.94-0.98, p< .001). More importantly, the results calculated using different sets of parameters are consistent and lead to the same conclusions. These three additional sets of parameters used to calculate the PLV are reported in Table A1 included in Supplemental Digital Content 1. GDTs plotted as a function for PLVs calculated using these additional parameters are shown in Figure A1 included in Supplemental Digital Content 2. The association between PLVs and CNC word scores measured in different conditions and the association between the PLV and the noise effect on CNC word scores are shown in Figures B1 and C1 included in supplemental Digital Content 3 and 4, respectively. The results of Pearson-Moment Product correlation tests are also shown in these two figures. Descriptive results of PLVs calculated using these three additional sets of parameters and LMMs results showing the effect of stimulation level and electrode location on the PLV are reported in Table B1 included in Supplemental Digital Content 5. Results of pairwise comparisons for comparing PLVs measured at different electrode locations are reported in Table C1 included in Supplemental Digital Content 6. Overall, these data suggest that this method does not rely on the specific parameters used in the time-frequency decomposition.

Second, the recording window for measuring the eCAP should not be confused with the frame size in time used in the time-frequency decomposition. The recording window offered by different CI manufacturers’ software ranges from 1561.6-2500 μs, which is longer than a time window where the eCAP is expected for in human CI users (i.e., within the first 1200 μs) (Botros et al., 2007). The inter-trial phase coherence is low for the part where the recorded traces only contain noise. As a result, including prolonged time window in the time-frequency decomposition is not always beneficial or appropriate. Admittedly, the duration of the recording window affects the number of frames that could be included in the time-frequency decomposition. However, our results reported in Supplemental Digital Contents suggest that it is not a determing factor for this method.

Third, Advanced Bionics and MedEL devices offer higher sampling rates (Advanced Bionics: 56 kHz, MedEL: 1.2 MHz) than Cochlear™ Nucleus® device for eCAP recording. This parameter is determined by each CI manufacturer and cannot be changed in their clinical software. In human CI users, the shortest possible interpeak latency of the eCAP is around 0.2 ms (for a review, see He et al., 2017). In a Fast Fourier Transform analysis, this corresponds to a fundamental period of 0.4 ms or a fundamental frequency of 2.5 kHz. Cochlear™ Nucleus® device offers a sampling rate of 20,492 Hz, and therefore the fundamental frequency and first few harmonics of these shortest eCAP waveforms fall below the Nyquist frequency of 10,246 Hz. Thus, this sampling rate, which is the lowest among all three major CI devices, is sufficient for eCAP recording and is not a factor that could limit the sensitivity of PLV measures. Nevertheless, the difference in sampling rate affects two parameters used in the time-frequency decomposition: the number of frames that can be included in the time-frequency decomposition and the number of samples that can be included in each frame. Higher sampling rates allow for shorter onset-to-onset intervals between two adjacent frames and more samples included in each frame. As a result, more frames with more samples included per frame can be included to calculate the PLV for Advanced Bionics and MedEL devices than for Cochlear™ Nucleus® device, which will affect the resulting PLV. This issue would not be problematic when comparing PLVs measured using the device from the same manufacturer. Only including the PLVs calculated for the frames with similar central spectral time windows to calculate the averaged PLV could eliminate the cross-device difference in the number of frames used in PLV calculation. One potential solution to minimize the effect of hardware-related difference on study results when comparing PLVs across devices is to normalize PLVs measured in individual participants based on the PLV range measured in a large group of patients with the same device, and to use the normalized results in comparison. This topic requires additional studies and is beyond the scope of this work.

Fourth, parameter modifications might be needed when applying this method to some patient populations whose morphological characteristics of the eCAP differ from those recorded in “typical” CI users. For example, eCAPs recorded in children with cochlear nerve deficiency show prolonged interpeak latencies and less prominent P2 peak, which could affect the frequency range used in the time-frequency decomposition.

Finally, results of previously published studies showed that the variation in discharge synchronization increases with pulse phase duration (Bruce et al., 1999) and during refractory recovery (Miller et al., 2001). The potential difference in spike initiation site for cathodic vs anodic stimulation also affects temporal jitter (Javel & Shepherd, 2000). As a result, the PLV could be affected by characteristics of the electrical pulse and the artifact rejection technique used to measured eCAP traces.

### Potential Study Limitations

This study has five potential limitations. First, this method is developed based on morphological characteristics of the eCAP measured in human CI users. Parameters used in this method may need to be modified to suit different animal models due to the anatomical difference in the CN between animal models and human listeners and the difference in durations and etiologies of deafness between experimental animals and human CI users. Second, the stimulation level could be a confounding factor for the results of this study because results of previous studies suggested a CN-health-dependent effect of stimulation level on peripheral neural synchrony. Specifically, Harris et al. (2021) reported larger PLVs measured at higher stimulation levels in listeners with acoustic hearing. However, this association was only observed in listeners with good CN health (i.e., young hearing listeners) and not in elderly listeners who have been shown to have poor peripheral neural synchrony and reduced CN densities. These results suggested that the stimulation level could be a confounding factor for the results of this study and thereby needs to be controlled for. Using stimulation levels that are balanced based on subjective perception of loudness has been widely used to control/minimize the potential stimulation level effect in psychophysical studies. In this study, both single-pulse and pulse-train stimuli delivered to different CI electrodes tested in each participant were presented at the levels that were determined to be “maximal comfort” (rating 8 on the same visual loudness rating scale). Therefore, in a certain sense and to a certain degree, the stimulation levels used at different electrodes can be considered loudness balanced within each participant and even across participants. However, electrical biphasic pulses that evoke neural responses with comparable amplitudes or these matched in current level are not necessarily perceived as equally loud by CI users (Kirby et al., 2012). Similarly, neural responses evoked by pulses that are perceived equally loud by CI users can have a large difference in neural response amplitude (Kirby et al., 2012). In addition, comparing loudness for single pulse stimulation with a total duration of only 57 µs could be impossible for many CI users, as demonstrated during our pilot study. Therefore, using loudness-balanced stimulation levels may not be a good solution to eliminate the potential level effect on study results. Instead, the stimulation level effect on results of this study was controlled using statistical analyses. Third, as an initial step toward understanding the role of peripheral neural synchrony in determining CI clinical outcomes, this study only evaluated the association between neural synchrony in the CN and monoaural auditory perception outcomes in human CI users. The modeling study by Resnick and Rubinstein (2021) suggests that degraded neural synchrony might have an even greater impact in binaural listening conditions where interaural timing difference cues are needed to help separate a speech stream from background noise. Further studies are warranted to determine the role of neural synchrony in the CN in binaural hearing. Fourth, despite these results, the exact biological underpinning (e.g., demyelination, peripheral axon degeneration, or total SGN loss) of the PLV remains unknown and requires further investigation. Due to the lack of noninvasive tools, it is not feasible to use experimental approaches to determine physiological conditions of the CN in living human listeners. Computational modeling techniques have been widely used to probe neuroanatomical conditions underlying neural response patterns in neuroscience, which holds the potential to be used to answer this important question. Finally, participants tested in this study generally showed better/higher CNC word scores than those previously reported (e.g., Bierer et al., 2016; Holder et al., 2020). Further studies in CI users with varied speech perception outcomes are warranted to fully assess the role of peripheral neural synchrony in determining auditory perception outcomes in electrical hearing.

## CONCLUSIONS

Neural synchrony in the electrically stimulated CN could be estimated at individual electrode locations in CI users by calculating the phase coherence across repeated presentations of a single pulse stimulus using the method reported in this paper. Peripheral neural synchrony varies across CI users and electrode locations. Poorer peripheral neural synchrony leads to lower temporal resolution acuity. The degree of peripheral neural synchrony is associated with the size of detrimental effect of competing background noise on speech perception performance in post-lingually deafened adult CI users. Further studies are warranted to fully understand the role of peripheral neural synchrony in determining auditory perception outcomes in electrical hearing.

## Supporting information

Supplemental Digital Content 1

Supplemental Digital Content 2

Supplemental Digital Content 3

Supplemental Digital Content 4

Supplemental Digital Content 5

Supplemental Digital Content 6

## Data Availability

Please contact the corresponding author to discuss access to the data presented in this study.

## Author Contributions

SH designed this study, participated in data analysis, drafted and approved the final version of this paper. JS participated in data collection and data analysis, provided critical comments, and approved the final version of this paper. IB participated in study design, provided critical comments, and approved the final version of this paper. JO conducted statistical analyses, provide critical comments, and approved the final version of this paper. YY participated in data collection, provided critical comments, and approved the final version of this paper.

