## Supplemental Digital Content 1 for "Peripheral neural synchrony in post-lingually deafened adult cochlear implant users"

**Table A 1. Three additional sets of parameters used to calculate the phase locking value (PLV)**

| Parameter Set | Frequency Range (Hz) | Number of Linearly Spaced Frequencies | Number of Samples Included in Each Frame | Frame Size in Time (μs) | Number of Frames Used for Calculating the Averaged PLV | The Time Window Used for Calculating the Averaged PLV (μs) |
|---|---|---|---|---|---|---|
| I | 1280.8 – 3201.9 | 4 | 16 | 780.8 | 6 | 179 – 1203.8 |
| II | 1280.8 – 4482.6 | 6 | 16 | 780.8 | 6 | 179 – 1203.8 |
| III | 683.1– 4781.5 | 13 | 30 | 1464 | 2 | 179 – 1561.6 |
