## Supplemental Digital Content 2 for "Peripheral neural synchrony in post-lingually deafened adult cochlear implant users"

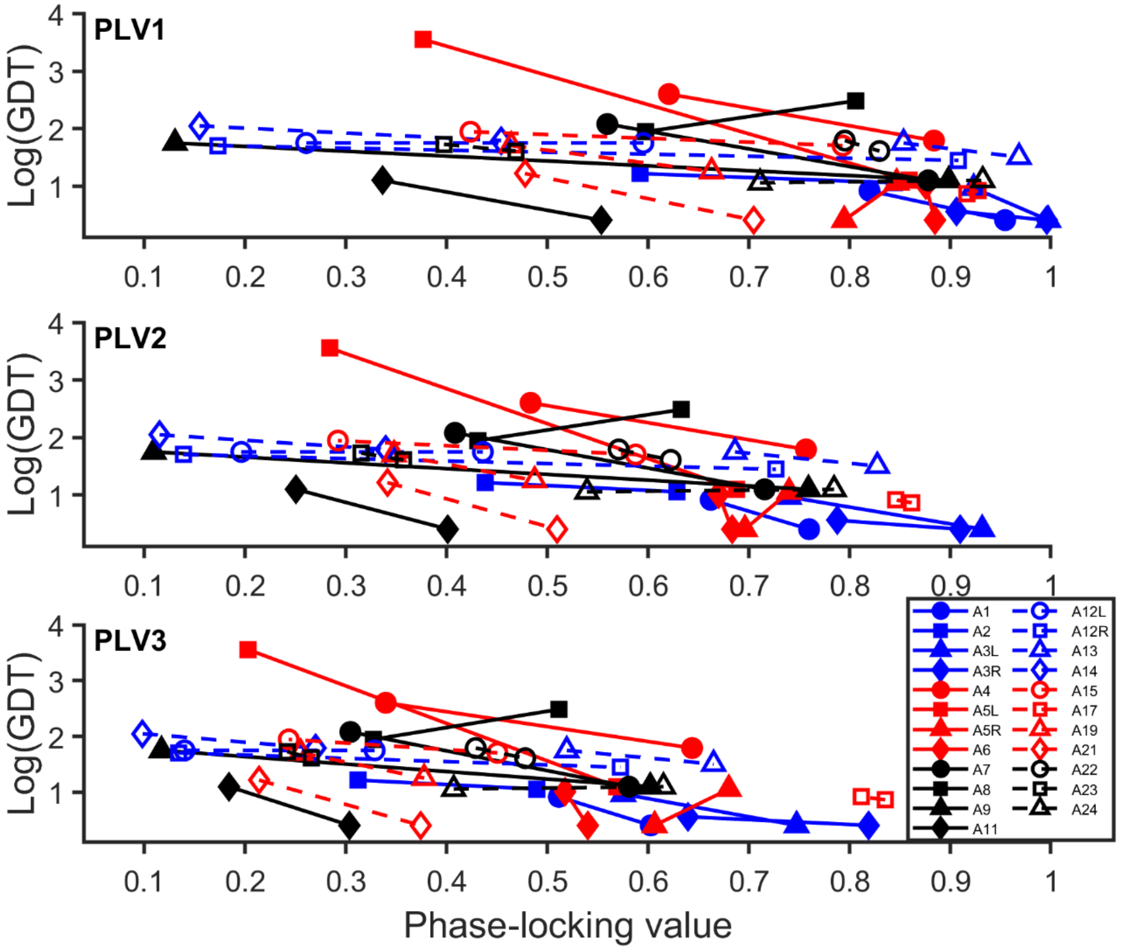


**Figure A 1.** Phase-locking values calculated using different sets of parameters (PLV1, PLVs calculated using Parameter set I; PLV2, PLVs calculated using Parameter set II; PLV3, PLVs calculated using Parameter set III) and psychophysical gap detection thresholds measured at two electrode locations in each of 23 implanted ears of 20 participants. Lines connect the data measured at the two electrode locations tested in the same ear.
