## Supplemental Digital Content 3 for "Peripheral neural synchrony in post-lingually deafened adult cochlear implant users"

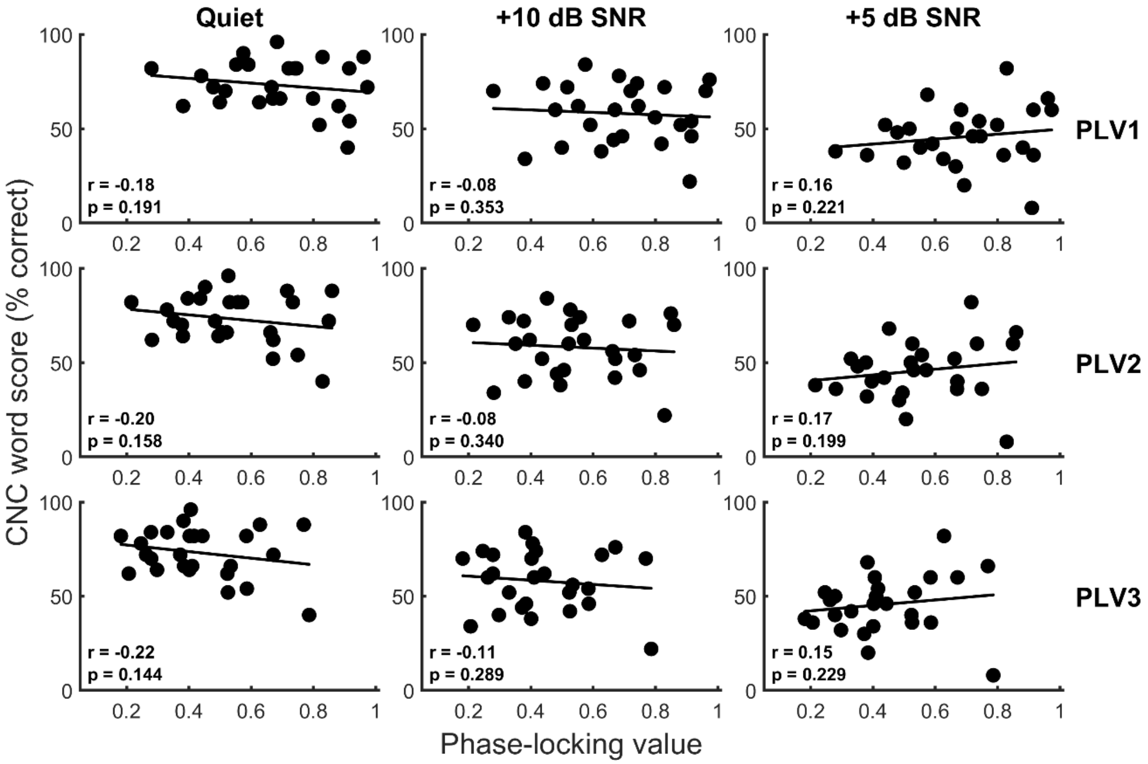


**Figure B1**. Consonant-Nucleus-Consonant (CNC) word scores measured in quiet and in two noise conditions as a function of the averaged phase-locking value (PLV) across electrode locations for 23 adult cochlear implant users (26 ears). PLVs calculated using different sets of parameters (PLV1, PLVs calculated using Parameter set I; PLV2, PLVs calculated using Parameter set II; PLV3, PLVs calculated using Parameter set III) are indicated in different rows. Each column indicates results measured in each testing condition. The best fit line across all 26 data points is illustrated with a solid line. The results from Pearson's correlation analysis are also provided in each panel.
