## Supplemental Digital Content 4 for "Peripheral neural synchrony in post-lingually deafened adult cochlear implant users"

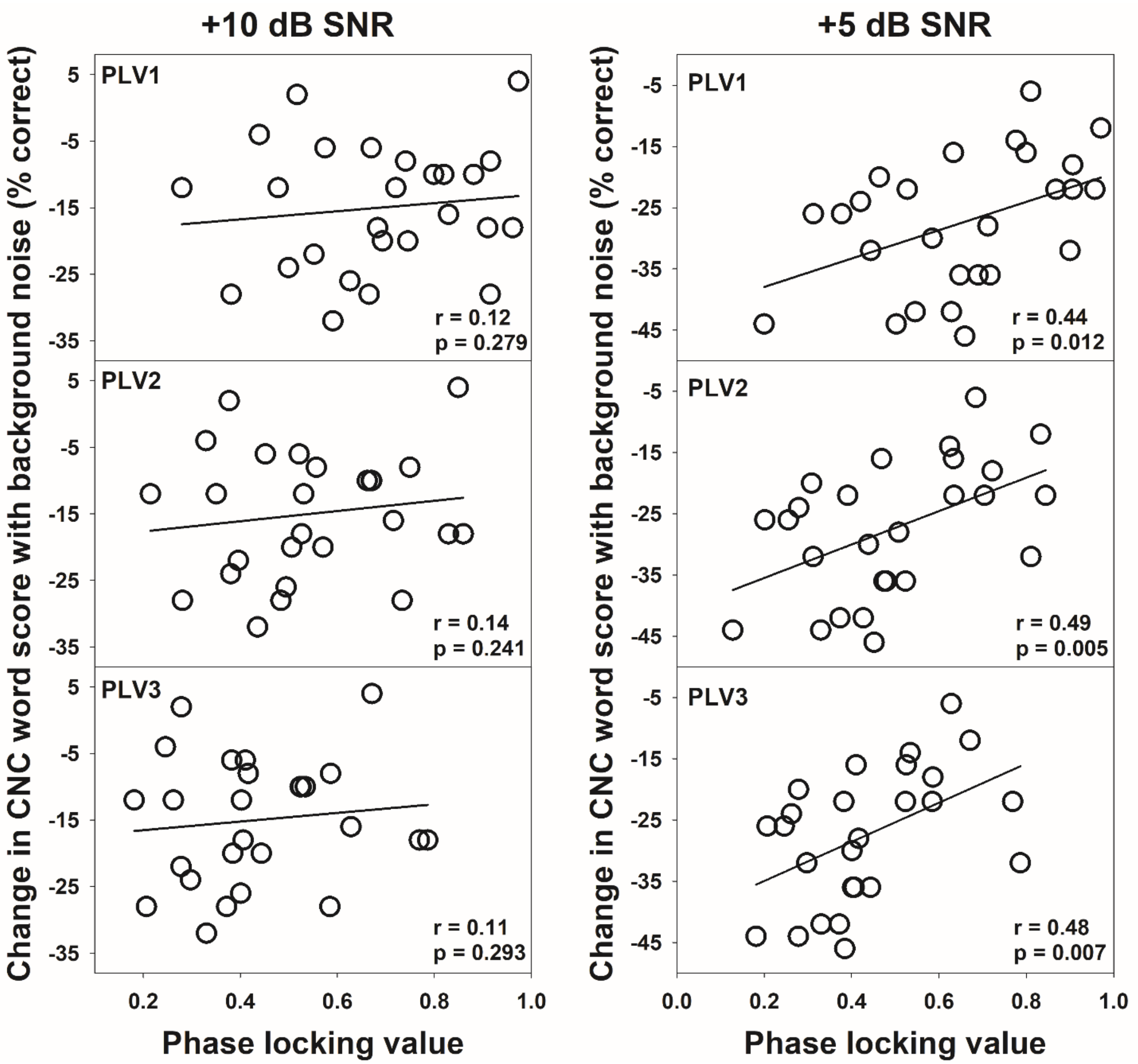


**Figure C1.** The change in Consonant-Nucleus-Consonant (CNC) word scores with the addition of background noise as a function of the averaged phase-locking value (PLV) across electrode locations. PLVs calculated using different sets of parameters are indicated in different rows (PLV1, PLVs calculated using Parameter set I; PLV2, PLVs calculated using Parameter set II; PLV3, PLVs calculated using Parameter set III). Each column indicates results measured in each testing condition. The best fit line across all

26 data points is illustrated with a solid line. The results from Pearson's correlation analysis are also provided in each panel.
