## Supplemental Digital Content 5 for "Peripheral neural synchrony in post-lingually deafened adult cochlear implant users"

**Table B 1. Results for phase locking values (PLVs) calculated using three additional sets of parameters.** PLV1, PLVs calculated using Parameter set I; PLV2, PLVs calculated using Parameter set II; PLV3, PLVs calculated using Parameter set III; STD, standard deviation; GDT, gap detection threshold.

| | Range (mean, STD) | Electrode Effect | Level Effect | Effect on GDT |
|---|---|---|---|---|
| PLV1 | 0.13 – 1.0 (0.69, 0.25) | $\chi^2_{(3)}$ = 14.72; p < .01 | $t_{(101)}$ = 2.30; p = .02 | $t_{(42)}$ = -3.20; p < .01 |
| PLV2 | 0.11 – 0.93 (0.55, 0.23) | $\chi^2_{(3)}$ = 15.58; p < .01 | $t_{(101)}$ = 3.07; p < .01 | $t_{(42)}$ = -3.25; p < .01 |
| PLV3 | 0.10 – 0.84 (0.44, 0.20) | $\chi^2_{(3)}$ = 15.07; p < .01 | $t_{(101)}$ = 3.49; p < .01 | $t_{(42)}$ = -3.60; p < .01 |
