## Supplemental Digital Content 6 for "Peripheral neural synchrony in post-lingually deafened adult cochlear implant users"

**Table C1. Results of pairwise comparisons for comparing phase locking values (PLVs) measured at different electrode locations.** PLV1, PLVs calculated using Parameter set I; PLV2, PLVs calculated using Parameter set II; PLV3, PLVs calculated using Parameter set III.

| | Electrode Pair | Estimate | Standard Error | Degree of Freedom | T Ratio | *p* Value |
|---|---|---|---|---|---|---|
| PLV1 | E3 vs. E9 | -0.05 | 0.03 | 29.0 | -1.16 | .39 |
| | E3 vs. E15 | -0.13 | 0.04 | 30.3 | -3.48 | <.01 |
| | E3 vs. E21 | -0.12 | 0.05 | 40.0 | -2.46 | .08 |
| | E9 vs. E15 | -0.09 | 0.02 | 27.2 | -3.51 | <.01 |
| | E9 vs. E21 | -0.08 | 0.04 | 36.5 | -1.91 | .24 |
| | E15 vs. E21 | 0.00 | 0.03 | 30.8 | 0.24 | .99 |
| PLV2 | E3 vs. E9 | -0.03 | 0.02 | 30.0 | -1.42 | .50 |
| | E3 vs. E15 | -0.11 | 0.03 | 29.7 | -3.59 | .01 |
| | E3 vs. E21 | -0.11 | 0.04 | 37.5 | -2.59 | .06 |
| | E9 vs. E15 | -0.08 | 0.02 | 27.2 | -3.57 | .01 |
| | E9 vs. E21 | -0.08 | 0.04 | 35.4 | -2.15 | .16 |
| | E15 vs. E21 | 0.00 | 0.03 | 31.2 | 0.10 | 1.00 |
| PLV3 | E3 vs. E9 | -0.02 | 0.02 | 30.1 | -1.03 | .73 |
| | E3 vs. E15 | -0.08 | 0.02 | 28.2 | -3.38 | .01 |
| | E3 vs. E21 | -0.09 | 0.03 | 33.4 | -2.67 | .05 |
| | E9 vs. E15 | -0.06 | 0.02 | 26.6 | -3.37 | .01 |
| | E9 vs. E21 | -0.07 | 0.03 | 33.1 | -2.29 | .12 |
| | E15 vs. E21 | 0.00 | 0.02 | 30.9 | 0.22 | 1.00 |
